# Family planning self-care: from global frameworks to local meaning, perceptions, experiences and opportunities in Niger

**DOI:** 10.64898/2026.04.08.26350458

**Authors:** Jean Christophe Fotso, Elikem Togo, Dieudonne Bidashimwa, Olaitan Elihou Adje, Nouhou Abdoul Moumouni

**Author notes:** Corresponding author: (JCF).

## Abstract

Family planning (FP) self-care is a strategic pillar for advancing Universal Health Coverage (UHC) and mitigating health workforce shortages. However, a significant disconnect persists between global normative frameworks and local implementation realities. This study examines the local meanings, perceptions, and experiences of FP self-care in Niger to inform contextualized scale-up of self-care interventions. We employed a sequential mixed-methods design in the Niamey (urban) and Zinder (rural) regions of Niger. A quantitative household survey was conducted with 510 women and 357 men to assess fertility awareness, method preferences, and information-seeking behaviors. This was complemented by qualitative in-depth interviews with 36 women, 18 men, 12 healthcare providers, and 15 community leaders. Quantitative data were analyzed using descriptive statistics, while qualitative transcripts underwent iterative thematic analysis mapped to global self-care frameworks. “Self-care” was locally reconstructed not as autonomy. While defined by all participants as hygiene, it was uniquely reconstructed by men and community leaders as economic provision. A distinct “medicalization paradox” emerged: women defined self-care as the agency to seek clinical dependence, prioritizing facility-based providers over community sources (e.g., 58.1% vs. 12.1% for oral contraceptives) to mitigate fears regarding product quality and side effects. Conversely, men favored Community Health Workers (34.3%) driven by logistical efficiency and economic motivations. Physiological knowledge was low; only 11.8% of women correctly identified the fertile window, with misconceptions reinforced by fatalistic narratives propagated by community gatekeepers. Furthermore, providers expressed strong skepticism regarding user competence, fearing “chaos” without medical supervision. Implementing FP self-care in Niger requires shifting from a “product-first” to a “values-first” approach. Strategies must be gender-stratified: leveraging “medicalized validation” to address women’s safety concerns while utilizing community-based channels to meet men’s efficiency needs. Ultimately, self-care should be framed not as independence from the health system, but as an empowered partnership with it.

## Introduction

Comprehensive sexual and reproductive health and rights (SRHR) is fundamental for Universal Health Coverage (UHC), yet achieving this remains persistently challenging [1]. With conventional health systems severely strained by humanitarian emergencies and pandemics, alongside an anticipated global deficit of up to 18 million health workers by 2030 [1,2], stakeholders recognize the urgent need for innovative, cost-effective strategies that shift away from facility-centric models to enhance access, maximize user autonomy, and broaden reproductive choice, thereby strategically advancing UHC goals [2–4].

The World Health Organization (WHO) defines self-care as the capacity of individuals to maintain health either autonomously or with health worker support [1,5]. Self-care interventions (SCIs) are evidence-based tools, including products, devices, diagnostics, and digital health applications, that support this practice [1]. This strategy, which includes high-relevance modalities such as self-administered injectable contraception (DMPA-SC SI) and OTC (Over-The-Counter) access to oral contraceptive pills (OCPs) [6], is underpinned by WHO consolidated guidelines, which mandate that scaling SCIs be predicated on human rights and gender equality principles and facilitated by a supportive environment that protects users against coercion, stigma, and discrimination [1,7].

The concept of family planning (FP) self-care is not new. Historically, the international FP community has acted as a vanguard in shifting access away from static facilities, evidenced by the introduction of checklists in the 1970s to facilitate community-based distribution (CBD), the scaling of social marketing initiatives, and long-standing advocacy for OTC purchase of oral contraceptives [1,8,9]. While contraceptive self-care is sometimes equated with self-injectable methods, the concept is theoretically broader, encompassing the acquisition of health literacy and the ability to use, when preferred, methods that require little or no advice from providers, such as fertility-awareness methods and condoms [1,7,8,10].

### Global Framing and Evidence Base on Self-Care

The global integration of SRHR SCIs is primarily guided by conceptual frameworks emphasizing people-centered and health systems approaches [2]. The People-Centered Approach focuses on empowering individuals by supporting their agency, autonomy, and self-determination, ensuring informed decisions regarding their SRHR [1,11]. Complementarily, the Health Systems Approach positions SCIs as strategies to optimize Primary Health Care (PHC) resources and efficiency [1,2]. Integrating SCIs must be recognized as a strategic investment rather than merely a cost-cutting measure. While self-care may reduce user costs, sustainable financing is necessary to support requisite system reforms and regulatory oversight [3].

Empirical evidence confirms the benefits of key SCIs in enhancing Reproductive Empowerment (RE) and individual agency [6]. Interventions such as OCPs, emergency contraception, and DMPA-SC (Depot Medroxyprogesterone Acetate Subcutaneous) offer cost-effective pathways to empowerment and privacy [1,6]. However, realizing these benefits is frequently challenged by power dynamics and reproductive coercion, where partners may manipulate or pressure individuals into sex without any form of FP [3,6].

The successful integration of SCIs must adhere to human rights standards to mitigate potential harm [7]. Yet, a significant disconnect persists regarding the translation of frameworks into local implementation strategies [10–12]. Effective uptake is intrinsically shaped by complex contextual factors spanning individual agency and systemic accessibility [10,13]. Therefore, qualitative inquiry is critically needed to explore the local meanings, preferences, and feasibility of self-care interventions [12,14].

This manuscript addresses this deficit by focusing on FP self-care in Niger, a context typified not only by resource limitations but by specific sociopolitical dynamics that may influence trust in non-clinical distribution. While specific methods like diaphragms and LAM (Lactational Amenorrhea Method) have been used [15], little is known about how the standardized concept of “self-care” is interpreted. The core research problem is the lack of localized evidence required to operationalize global recommendations [3,16]. Translating normative guidance into effective national policies demands robust local implementation research [13]. Consequently, this study aims to contextualize local experiences of FP self-care in Niger within current global frameworks. Specifically, we seek to: 1) Examine the meaning, perceptions, and experience with contraceptive self-care; and 2) Explore opportunities to improve SRHR.

The insights generated will be paramount for guiding the equitable scale-up of self-care strategies, informing the adaptation of guidelines essential for accelerating UHC progress in Niger [2,3].

## Methods

This paper draws upon data collected in Niger under the multi-country Research for Scalable Solutions (R4S) project, which also included Uganda and Nepal [17]. To operationalize the study objectives, we employed a sequential mixed-methods design, integrating a quantitative household survey with nested qualitative in-depth interviews (IDIs). This design was critically selected to not only quantify awareness and behaviors but to qualitatively unpack the specific sociocultural drivers behind observed uptake patterns. Study sites in Niger were purposively selected to reflect distinct sociocultural and geographical contexts: the urban Niamey region and the predominantly rural Zinder region. Data collection instruments were structured to assess adherence to WHO guidelines across three key FP stages: awareness, access, and use. All data were collected in the dominant local languages: French, Haoussa, and Zerma.

### Quantitative component: Household survey

The survey utilized a two-stage stratified cluster sampling design. Primary Sampling Units (PSUs), based on census enumeration areas, were stratified by urban/rural classification. From this frame, 25 PSUs were selected using probability proportionate to size (PPS). Subsequently, a comprehensive household listing was performed within these PSUs to identify households with at least one woman of reproductive age. Twenty-four households were then randomly selected per PSU, creating a sample frame of 600 households. The study aimed for a final sample of 500 women and 250 men. Eligible women were aged 15–49, married or in a union within the previous three months, and were either current contraceptive users or had an unmet need for family planning. In approximately half of the sampled households, eligible men, aged 18–59, in a current relationship, and not sterilized, were invited to participate.

The survey instrument gathered data on socio-demographics, contraceptive use history, fertility awareness, access to FP information, including interest in digital health platforms, method awareness, and perceived importance of provider/Community Health Worker (CHW) engagement in FP access and use. Specific self-care modules assessed preferences for obtaining and managing side effects for OCPs, emergency contraception (EC), and DMPA-SC self-injection (DMPA-SC SI). Current or past contraceptive users were additionally asked about method choice and source of supply. Data was collected via ODK (Open Data Kit)-programmed mobile devices.

### Qualitative component: In-depth interviews (IDIs)

To explain the quantitative patterns, qualitative participants were nested within the survey PSUs. A purposive sampling strategy was employed to select participants from a subset of surveyed households and to ensure variation in age, marital status, and contraceptive experience. The study targeted 36 women (one or two women per PSU) and 18 men (one in half of the households selected for women IDIs). To capture systemic perspectives, we recruited key informants, including 12 healthcare providers (public, private, and pharmacy sectors) and 15 community leaders, maintaining an urban-rural balance.

The IDIs aimed to provide a contextual understanding of self-care, focusing on perceptions of and experiences with contraceptive self-care behaviors, as well as barriers and facilitators to FP practice. The interviews explored the role(s) that women, men, and providers play in accessing and managing FP information, methods, and services. To capture authentic local meanings without bias, interviews began with open-ended questions regarding the general meaning of “self-care” before focusing on family planning. If respondents found the term difficult to conceptualize, a study-specific adaptation of the World Health Organization (WHO) definition was provided as “the ability of individuals, families, or communities to promote and maintain sexual health and avoid unintended pregnancies with or without the help of a healthcare provider”. All sessions were audio-recorded and subsequently transcribed into French for analysis.

### Analysis methods

Descriptive analysis of the survey data was conducted using STATA version 15. To account for the multi-stage sampling design, analyses incorporated sampling weights and adjusted for clustering effects. Qualitative transcripts were managed and analyzed using NVivo 12 and NVivo 20 (Release 1). The research team followed a rigorous, iterative thematic analysis process:

1. Codebook Development: A comprehensive codebook was developed and applied consistently across all sites.
2. Familiarization and Quality Check: Transcripts underwent thorough quality checks and initial reading for thematic immersion.
3. Thematic Analysis: Thematic memos and matrices were utilized to systematically assess content differences across geography, gender, and stakeholder type.
4. Data Synthesis: The final stage involved linking localized findings on meaning, perceptions and opportunities back to the global self-care frameworks established in the literature review.

### Ethical considerations

The study was approved by the FHI (Family Health International) 360 Office of International Research Ethics and Niger’s Comité National d’Ethique pour la Recherche en Santé (CNERS). Participation was voluntary; women provided verbal consent for eligibility screening, and all eligible participants (women and men in the survey and all IDI participants) provided written informed consent prior to data collection.

## Results

### Sample characteristics

**Table 1** presents the background and contraceptive profiles of survey respondents (510 women and 357 men). The sample is characterized by high marriage rates (97.9% women; 96.4% men) and low educational attainment, with 61.5% of both women and men lacking formal schooling. A significant age and employment disparity exists: 57.8% of women are under 35 years, compared to only 30.8% of men, and women are significantly less likely to have engaged in recent paid work (23.4% vs. 39.6%). While geographical access is high - 74.1% of women reside within 30 minutes of a facility - modern contraceptive prevalence remains at 30.9%, with the method mix heavily skewed toward short-acting options like pills (53.7%) and injectables (28.6%). Fertility intentions primarily favor birth spacing; however, a striking gender divergence appears regarding birth limiting: 14.5% of women wish to stop childbearing entirely, compared to only 2.5% of men.

**Table 1.** Background characteristics and contraceptive use history.

|  | Women (n=510) |  | Men (n=357) |  |
| --- | --- | --- | --- | --- |
|  | N | % | N | % |
| **Residence/site** |  |  |  |  |
| Urban (largely Niamey)<sup>1</sup> | 293 | 53.7 | 154 | 36.7 |
| Rural/Zinder | 217 | 46.3 | 203 | 63.3 |
| **Marital Status** |  |  |  |  |
| Married | 500 | 97.9 | 339 | 96.4 |
|  | N | % | N | % |
| Other (divorced/widowed, never married) | 10 | 2.1 | 18 | 3.6 |
| **Education Level** |  |  |  |  |
| No formal schooling | 296 | 61.5 | 215 | 61.5 |
| Primary school | 91 | 16.4 | 58 | 19.1 |
| Secondary school+ | 123 | 22.1 | 84 | 19.4 |
| **Age** |  |  |  |  |
| < 24 | 113 | 19.8 | 22 | 5.5 |
| 25 - 34 | 185 | 38 | 94 | 25.3 |
| 35+ | 212 | 42.2 | 241 | 69.2 |
| **Had Paid Work in Last 6 Months** |  |  |  |  |
| Yes | 126 | 23.4 | 133 | 39.6 |
| No | 384 | 76.6 | 224 | 60.4 |
| **Travel Time to Nearest Preferred Health Facility** |  |  |  |  |
| Less than 15 minutes | 195 | 38.7 | 206 | 61.5 |
| 15 to 30 minutes | 174 | 35.4 | 88 | 22 |
| 31-60 minutes | 79 | 12.8 | 38 | 10.7 |
| More than one hour | 42 | 7.7 | 7 | 1.8 |
| Don't know | 20 | 5.4 | 18 | 4 |
| **Modern contraceptive use status** |  |  |  |  |
| Current users of modern contraception | 175 | 30.9 | 68 | 17.7 |
| Non-users that are past users of modern contraception | 105 | 20.7 | 25 | 6.1 |
| Never users of modern contraception | 230 | 48.4 | 264 | 76.2 |
| **Method mix<sup>2</sup>** |  |  |  |  |
| Implant | 38 | 13.4 | 6 | 7.6 |
| IUD | 11 | 3.6 | 4 | 2.7 |
| Injectable | 82 | 28.6 | 25 | 27.4 |
|  | N | % | N | % |
| Pill | 146 | 53.7 | 44 | 45.8 |
| LAM & Male condom | 3 | 0.7 | 14 | 16.5 |
| **Fertility Intentions<sup>3</sup>** |  |  |  |  |
| I want a child within the next 2 years |  |  | 137 | 45.3 |
| Want to wait 2+ years before the birth of a child | 136 | 43.9 | 63 | 29.7 |
| Want a child but don't know when | 109 | 33.1 | 56 | 21.9 |
| Don't know if wants a child | 28 | 8.5 | 4 | 0.6 |
| Don't want any or any more children | 51 | 14.5 | 13 | 2.5 |

<sup>1</sup>A few EAs in Zinder were also urban;
<sup>2</sup>Among current and past users of modern methods (n=280);
<sup>3</sup>Among women (n=324) and men (n=273) who answered this question

### Meaning of self-care

Qualitative inquiries revealed that the global concept of “self-care” is locally reconstructed through three distinct lenses: hygiene, economic provision, and clinical agency. Universally, participants defined self-care as the maintenance of personal and environmental hygiene. However, a gendered economic dimension emerged among men and community leaders, who explicitly linked “care” to financial survival rather than health maintenance. A leader from Zinder articulated this provision-based definition: *“Doing everything possible to help your children with an economic activity… Through this activity, they can rely on their own abilities as actors”*. Male respondents echoed this, defining self-care as the agency derived from income generation: *“It is when I have an activity that allows me to take care of my problems”*.

In the specific context of FP, the concept proved difficult for participants to operationalize, revealing a sharp dichotomy between ‘autonomous use’ and ‘responsible consultation’. While a minority favoring traditional methods defined self-care as managing fertility *“without resorting to health workers”*, the dominant interpretation - shared by women and providers - paradoxically defined self-care as the active effort to seek professional dependence. Participants argued that true agency necessitates clinical supervision. A provider summarized this view: *“Self-care in FP cannot be an option that is left to clients without going through a health worker”*. This opinion corroborated by women who viewed the clinic visit not as a surrender of autonomy, but as a responsible act of *“taking charge of yourself”*

A minority counter-narrative existed, primarily among those favoring traditional methods, who defined self-care as managing fertility *“without resorting to health workers by using natural contraceptive methods”.* A health provider summarized this non-clinical perspective: *“It is about a woman taking charge of her contraception without resorting to health workers by using natural contraceptive methods… for example, the LAM method”*.

### Fertility Awareness and Information-Seeking Behaviors

Results in **Table 2** on fertility awareness expose critical knowledge deficits that fuel fatalistic attitudes. While general knowledge of menarche and cycle duration was high, precise identification of the fertile window was exceptionally low: only 11.8% of women and 2.9% of men correctly identified the mid-cycle period as the peak fertility window. Instead, a plurality (43.4% of women; 51.8% of men) adhered to vague beliefs that pregnancy risk exists broadly across ‘5 to 7 days’, failing to pinpoint ovulation.

**Table 2.** Fertility awareness, access to FP information, and interest in mobile access to FP information.

|  | Women (n=510) |  | Men (n=357) |  |
| --- | --- | --- | --- | --- |
|  | N | % | N | % |
| **Fertility awareness - participants who correctly know that:** |  |  |  |  |
| Girls typically start their menstrual cycle between 10 and 15 years old | 435 | 85.5 | 275 | 77.2 |
| The main sign that a girl is able to get pregnant is the first montly bleeding | 323 | 62.3 | 136 | 35.4 |
| The menstrual cycle starts on the first day of monthly bleeding | 348 | 66.8 | 273 | 75.4 |
| The menstrual cycle typically lasts between 26 and 32 days | 365 | 71.5 | 239 | 70.5 |
| Women are most likely to become pregnant halfway between the beginning of one period and the beginning of the next | 47 | 11.8 | 16 | 2.9 |
| A woman can get pregnant during 5 to 7 days per cycle | 210 | 43.4 | 195 | 51.8 |
| Sought information about FP and fertility in last 12 months<sup>1</sup> | 140 | 27.4 | 84 | 29.2 |
| **Sources used to seek information about FP and fertility in last 12 months<sup>2,†</sup>** |  |  |  |  |
| Clinic or health providers | 73 | 51.3 | 34 | 40.7 |
| Community Health Worker | 60 | 42.3 | 56 | 77.9 |
| Pharmacy/Drug Shop | 12 | 9.0 | 10 | 9.2 |
| Friend/Family Member | 21 | 14.4 | 6 | 4.5 |
| Digital sources - internet, websites, social media, voice or SMS messages | 1 | 1.7 | 3 | 3.3 |
| Radio or TV | 2 | 0.8 | 11 | 12.4 |
| **Interest in mobile access to FP information<sup>†</sup>** |  |  |  |  |
| Internet browser | 16 | 2.9 | 18 | 4.4 |
|  | N | % | N | % |
| Social media (WhatsApp, Facebook, Twitter, Instagram) | 79 | 13.5 | 58 | 15.2 |
| Voice/SMS message | 33 | 6.5 | 28 | 11.0 |
| Phone call | 145 | 27.4 | 74 | 25.0 |
| Health-related App | 29 | 5.8 | 43 | 13.0 |

<sup>1</sup>among all women, those who sought information on any of these topics: when a woman can get pregnant, contraceptive method choice, or management of contraceptive side effects;
<sup>2</sup>among women who sought information about FP in the 12 months before the survey;
<sup>†</sup>multiple responses were allowed

Qualitative data elucidate that these misconceptions are often diametrically opposed to biological reality. A prevalent error among women was the belief that the immediate post-menstrual period represented the highest risk, with one respondent stating, *“As soon as a woman finishes her period… her placenta is ready”*. This biological ignorance is compounded by a narrative of fatalism, particularly among men who rejected cycle tracking in favor of divine will. As one male respondent asserted: *“We can’t assume that we can get pregnant on a certain day because it’s something that comes from God”*. This external locus of control renders fertility-awareness methods perceived as futile.

Information-seeking behaviors reveal a distinct gender divergence regarding trust. While engagement levels were comparable, women relied primarily on clinical providers (51.3%) and Community Health Workers (CHWs) (42.3%), whereas men overwhelmingly utilized CHWs (77.9%). Qualitative findings clarify that women prioritize clinical encounters to validate safety against community misinformation, such as rumors of sterility or method migration. Conversely, men’s reliance on CHWs aligns with their exposure to community campaigns and preference for logistical efficiency.

Crucially, the “medicalization” sought by women serves as a strategic bypass of erroneous health advice circulated by community gatekeepers. Qualitative interviews revealed that influential leaders harbor specific fallacies, such as the belief cited by a female leader that oral contraceptives cause the uterus to contract, rendering future childbirth impossible. However, the clinical encounter itself is often imperfect; providers noted that women often arrive with fixed method preferences and limited motivation for counseling, suggesting that while women seek the *safety* of the clinic, technical inquiries often remain unvoiced. A significant discordance exists regarding the *quality* of information exchange within clinical settings. While women expressed a distinct desire for detailed technical information - specifically regarding product expiration, chemical composition, and side-effect management - providers conversely reported that users often arrive with fixed method preferences and limited motivation to seek counsel unless complications arise. This suggests that women’s technical questions often remain unvoiced during the encounter.

Finally, a “digital void” persists: despite latent interest in low-tech modalities like phone calls (27.4% of women), uptake of internet or app-based tools was negligible, underscoring that credibility remains rooted in interpersonal validation

### Preferences for Contraceptive Supply Sources

Preferences for supply sources expose a clear tension between logistical convenience and the imperative of clinical safety. As shown in **Table 3**, women exhibited a dominant preference for facility-based providers across all self-care modalities: 58.1% preferred clinicians for oral contraceptives versus 12.1% for CHWs, with similar trends for emergency contraception. Self-injection elicited the highest resistance, with nearly one-third of women explicitly stating they were “not interested,” irrespective of the source. In sharp contrast, men displayed a distinct preference for decentralized access, selecting CHWs (34.3%) significantly more often than clinical providers (22.4%). Qualitative data rationalize this divergence: men, who control family logistics but do not experience physiological side effects, prioritize the efficiency and accessibility of CHWs to minimize travel and waiting times. As one man noted, decentralized depots are preferable because they are *“easy and effective”*.

**Table 3.**
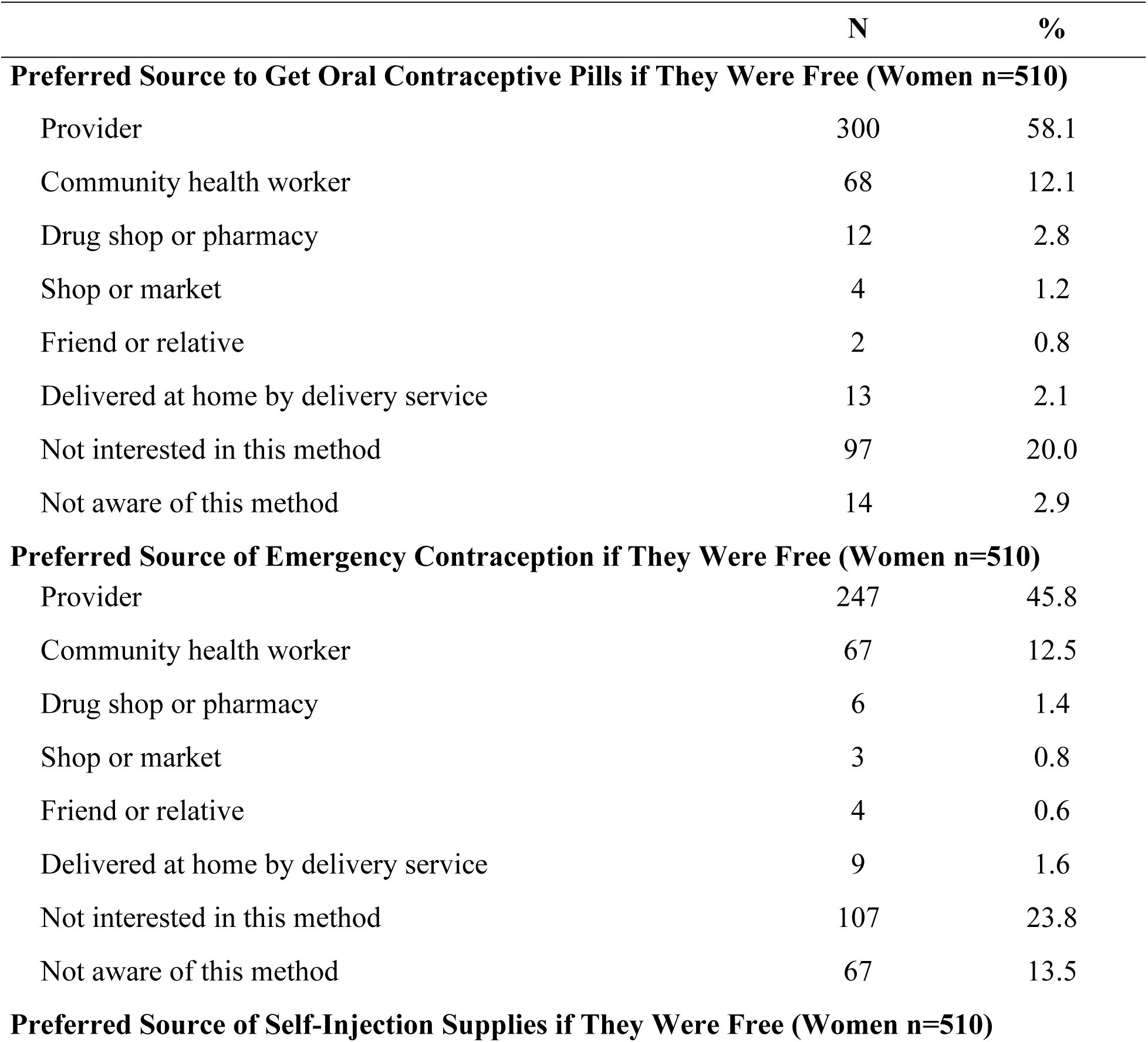
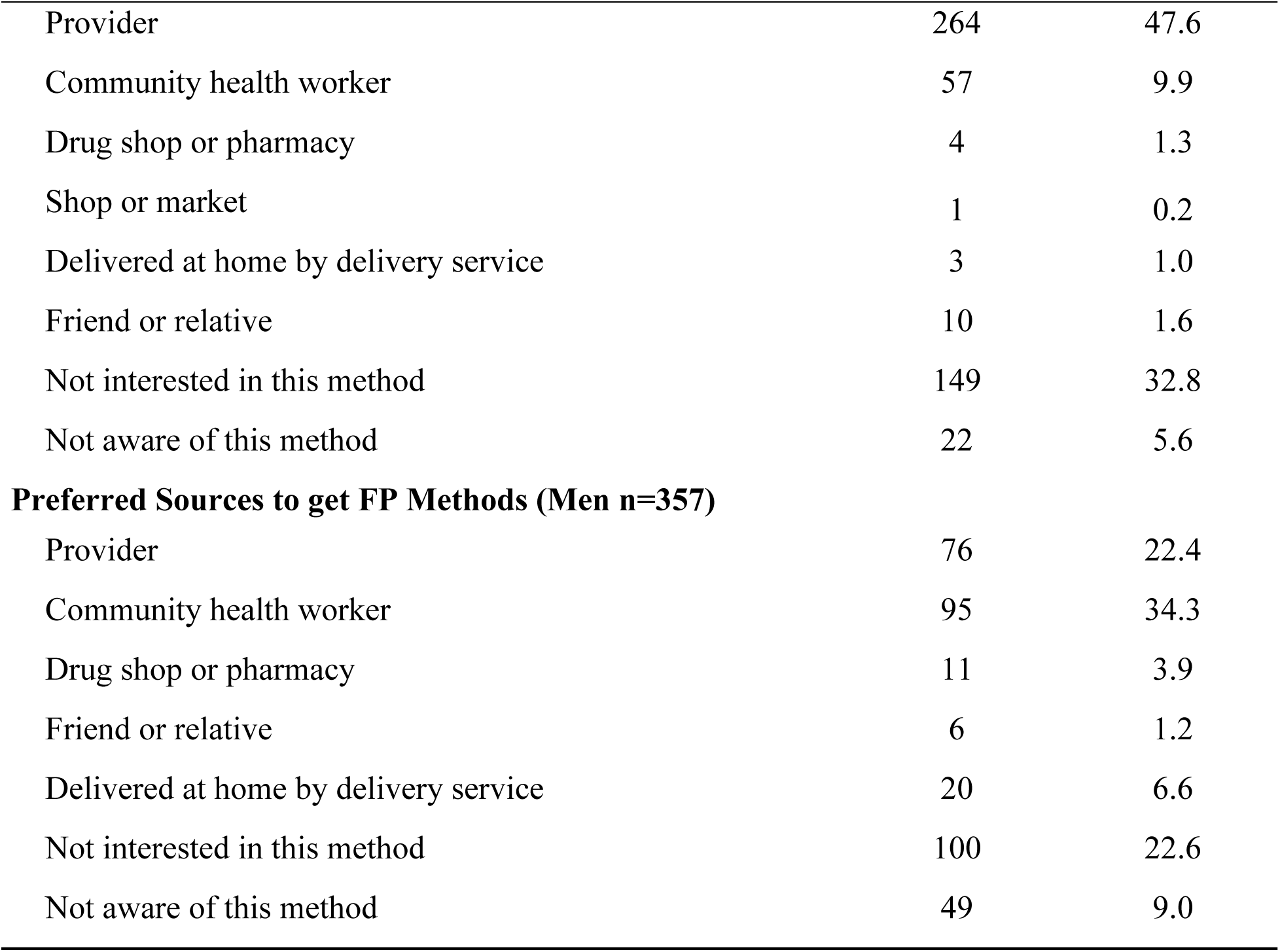
Preferences related to sources of supply for oral contraceptive pills, emergency contraception, and self-injection among all women and men.

|  | N | % |
| --- | --- | --- |
| **Preferred Source to Get Oral Contraceptive Pills if They Were Free (Women n=510)** |  |  |
| Provider | 300 | 58.1 |
| Community health worker | 68 | 12.1 |
| Drug shop or pharmacy | 12 | 2.8 |
| Shop or market | 4 | 1.2 |
| Friend or relative | 2 | 0.8 |
| Delivered at home by delivery service | 13 | 2.1 |
| Not interested in this method | 97 | 20.0 |
| Not aware of this method | 14 | 2.9 |
| **Preferred Source of Emergency Contraception if They Were Free (Women n=510)** |  |  |
| Provider | 247 | 45.8 |
| Community health worker | 67 | 12.5 |
| Drug shop or pharmacy | 6 | 1.4 |
| Shop or market | 3 | 0.8 |
| Friend or relative | 4 | 0.6 |
| Delivered at home by delivery service | 9 | 1.6 |
| Not interested in this method | 107 | 23.8 |
| Not aware of this method | 67 | 13.5 |
| **Preferred Source of Self-Injection Supplies if They Were Free (Women n=510)** |  |  |
| Provider | 264 | 47.6 |
| Community health worker | 57 | 9.9 |
| Drug shop or pharmacy | 4 | 1.3 |
| Shop or market | 1 | 0.2 |
| Delivered at home by delivery service | 3 | 1.0 |
| Friend or relative | 10 | 1.6 |
| Not interested in this method | 149 | 32.8 |
| Not aware of this method | 22 | 5.6 |
| **Preferred Sources to get FP Methods (Men n=357)** |  |  |
| Provider | 76 | 22.4 |
| Community health worker | 95 | 34.3 |
| Drug shop or pharmacy | 11 | 3.9 |
| Friend or relative | 6 | 1.2 |
| Delivered at home by delivery service | 20 | 6.6 |
| Not interested in this method | 100 | 22.6 |
| Not aware of this method | 49 | 9.0 |

For women, the preference for clinical settings acts as a rational risk-mitigation strategy against fears of product quality and bodily harm. Community-based sources are often viewed with suspicion due to rumors of expired or counterfeit stock. A sociopolitical dimension further complicates this, as community leaders described a “Nigerien mentality” of distrust toward free government distribution. A leader in Niamey explained: *“People often say that the products offered to them at home are not reliable… When the government does so, there is something fishy about it”*. Consequently, women seek clinical supervision to counter terrifying narratives, such as the fear that implants might *“disappear into the body”* or cause cancer.

Ultimately, the low interest in autonomous methods is reinforced by a provider-driven narrative regarding self-efficacy. While a minority of women expressed confidence in tasks like IUD self-removal, providers expressed strong opposition, predicting significant harm if medical supervision is removed. One provider vividly warned: *“When everyone is allowed to do whatever they want with contraception, there will be chaos… Self-medication is really not a good option”*. Thus, the data suggest that in this context, “autonomy” is frequently equated with danger, while safety is synonymous with clinical dependency.

While community leaders acknowledged the theoretical benefits of home-based care, they noted that the population views door-to-door delivery with suspicion, preferring the perceived legitimacy of the clinic. As one leader in Niamey explained: *“When you find a Nigerien with a product at home, he will doubt that product… He prefers to go out and find the product himself… People often say that the products offered to them at home are not reliable because, for them, the government is not generous. Consequently, the government cannot distribute something that is good door-to-door. When the government does so, there is something fishy about it”*.

Furthermore, clinical supervision is sought to counter terrifying community rumors regarding side effects. Women reported widespread narratives that implants could ‘disappear’ into the body or cause cancer. A woman from Zinder shared her hesitation: *“They said that it [the implant] disappears into the body… I’m really afraid of other methods. Implants, for example, have complications… She bled every time and even lost consciousness”*. Thus, the ‘medicalization’ observed in the survey reflects a risk-mitigation strategy where women prioritize physical safety and reassurance over convenience.

## Discussion

This study illuminates a distinct epistemological divergence between global normative frameworks of FP self-care and the lived realities of women, men, providers and community leaders in Niger. While international guidelines and literature typically frame self-care as a mechanism for enhancing autonomy, privacy, and reducing reliance on facility-based services [1,2], our findings reveal that the concept is locally reconstructed. In Niger, “self-care” is frequently interpreted as “compliance with hygiene” [18], or as “economic self-reliance” - a perspective explicitly articulated by community leaders who equate ‘care’ with the financial capacity to provide for one’s family. This local conceptualization stands in contrast to studies in other settings where self-care is primarily valued for its ability to bypass clinical gatekeepers [2,7].

A central finding of this study is the “medicalization paradox”. While the global self-care movement often emphasizes de-medicalization to demystify care and increase access [2,19], our data indicate that Nigerien women prioritize “medicalization” as a proxy for safety and validation. The strong preference for obtaining methods like oral contraceptives and injectables from clinical providers, rather than community sources, stems from deep-seated fears regarding product toxicity, expiration, and the ‘fishy’ nature of free government distribution noted by local leaders. Another study provides cross-national qualitative evidence confirming that fear of side effects, often based on rumors, is a dominant barrier, but that willingness to use increases significantly when a provider validates safety [20]. In a context where misinformation is not only rife but occasionally propagated by influential gatekeepers - such as the specific myths regarding uterine damage cited by female community leaders **-** the clinical encounter is viewed not as a barrier to autonomy, but as a necessary validation of it. This suggests that in settings with low health literacy and high prevalence of rumors, independent use without provider sanction is often viewed as risky or illegitimate rather than empowering.

Furthermore, our findings regarding source preferences highlight a significant gendered dichotomy relative to global trends. While broader literature demonstrates high acceptability of pharmacy-based access and digital health interventions for their convenience and anonymity [21], our study identifies a digital void and skepticism toward non-clinical sources among women. Conversely, the finding that men in Niger are more receptive to community-based distribution (e.g., CHWs) for logistical efficiency aligns with global evidence suggesting men value convenient, non-traditional entry points for SRH services to overcome structural barriers [22,23]. For men, who largely control the financial and permission-granting aspects of FP in this context, the “efficiency” of the source is paramount [24], whereas for women, who bear the physiological burden of side effects and the social risks of ‘unsupervised’ use, the “safety” of the source is non-negotiable.

Widespread apprehension over side effects and limited educational attainment in Niger drive a reliance on medicalized family planning (FP) services. This dependency is reinforced by low self-efficacy, positioning formal health workers as the primary gatekeepers of legitimate care. Research by Dougherty et al. [25] highlights behavioral heterogeneity within this population, suggesting that FP self-care is perceived differently across socio-economic strata. While less educated women may defer to clinical authority, more educated counterparts often demonstrate stronger behavioral agency. Additionally, implementation strategies remain critical in shaping women’s perception and self-efficacy. Even among women served by professional service providers, targeted counseling significantly correlates with higher perceived service quality and patient satisfaction [26], potentially mitigating the specific fears of product degradation cited by community stakeholders.

Finally, the relationship between knowledge, self-efficacy, and uptake in our study complicates standard empowerment frameworks. Global studies frequently posit that self-care interventions enhance reproductive empowerment by increasing body literacy and agency [6]. In contrast, our results expose critical deficits in foundational physiological knowledge—specifically the widespread misidentification of the fertile window, which fuels fatalistic attitudes attributing pregnancy solely to divine will [9]. Unlike studies where high self-efficacy facilitates the adoption of methods like self-injection [27], Nigerien providers expressed significant skepticism regarding client competence, fearing “chaos” without supervision. Consequently, reproductive empowerment in this context is less about the technical mastery of self-care tools and more closely tied to the negotiation of spousal permission, economic survival, and the ability to access quality care to counter the fatalism reinforced by community narratives.

### Opportunities for Advancing SRHR in Niger

The local reinterpretation of self-care does not negate the utility of the global framework; rather, it highlights the need for context-specific adaptation. Based on our findings, we identify four strategic opportunities to leverage self-care for improving SRHR in Niger.

#### Reconceptualizing “medicalization” as an enabler of trust and safety

The user preference for clinical providers should not be viewed as a failure of self-care adoption, but as an opportunity to implement a “supported self-care” model [28]. The WHO conceptual framework emphasizes that self-care exists on a continuum of interaction with the health system [1,2,27]. In Niger, where trust is anchored in the clinic, the opportunity lies in shifting the provider’s role from “gatekeeper” to “coach” [8,12]. Interventions should focus on equipping providers with competency-based training to transition from administering methods to educating clients on self-management [1]. Evidence suggests that when health workers personally understand or use self-care methods, their confidence in recommending them increases [12]. Programs should position self-care products (like DMPA-SC) not as a way to bypass the system, but as clinically validated tools that extend the clinic’s safety net into the home, thereby satisfying the user’s need for safety while reducing the frequency of facility visits [1].

#### Leveraging men’s efficiency motivations to expand community-based access

The distinct gender divergence in sourcing preferences—women preferring clinics for safety and men preferring CHWs for efficiency—presents a strategic entry point for male engagement, mostly in a context where husbands and community leaders are aware of their role on maternal health [29]. Given that men often hold decision-making power regarding FP in Niger, programs can leverage their preference for community-based distribution to overcome structural barriers such as distance and stock-outs [30]. By validating CHWs and depots as “efficient” sources for reputable products, programs can appeal to men’s role as resource providers [22,31]. Furthermore, since men in our study defined self-care partly through the lens of “economic autonomy”, FP advocacy in Niger should explicitly link contraceptive self-care to household economic stability and resilience. This framing moves FP away from purely “health” discourse toward “economic survival,” a definition that resonates with local male stakeholders [22]. This narrative, however, may attract criticism among policymakers who may view it as politically sensitive.

#### Addressing the “chaos” narrative through structured task-sharing

Provider skepticism regarding client capacity for self-care, described as a fear of “chaos,” mirrors global challenges where health workers view self-care as a threat to quality of care. Several studies in contexts provide empirical data needed to refute this “chaos” [31–34]. In the Niger context, the skepticism offers an opportunity to formalize “rational task-sharing” - a core component of an enabling environment for self-care [1]. To address provider resistance, implementation strategies must demonstrate that self-care is a mechanism for health system efficiency, not abandonment. The WHO guidelines highlight that self-care interventions can reduce the burden on overstretched health workforce [1]. The opportunity here is to introduce self-care protocols that include clear “safety valves” - robust referral mechanisms and decision-support tools - that reassure providers that users remain connected to the health system [1].

#### Mitigating fatalism through targeted fertility awareness

The critically low knowledge of the fertile window and prevalent fatalism identified in our study, a finding consistent with the 2021 ENAFEME Survey (Enquête Nationale sur la Fécondité et la Mortalité des Enfants de moins de cinq ans), which showed that “only 25% of women have a complete knowledge of the period when a woman has the greatest chance of becoming pregnant” [35], act as barriers to modern method uptake. However, the high local demand for “knowledge” and “hygiene” suggests an opportunity to reintroduce fertility awareness not merely as a contraceptive method, but as a fundamental component of “body literacy” and self-care [1]. Simple, low-tech self-care interventions can serve as entry points for broader SRH engagement. By demystifying the menstrual cycle, these tools can challenge fatalistic narratives and build the self-efficacy required for more complex contraceptive decision-making. Since digital literacy remains low among women in our sample, these interventions should rely on visual aids and interpersonal communication via CHWs rather than complex digital applications [36]. Correcting the widespread misconception that the post-menstrual phase is the most fertile period is a low-cost, high-impact self-care opportunity that requires minimal infrastructure but yields significant empowerment dividends [1].

### Limitations

This study is subject to a few limitations. While the stratification of study sites in Niamey and Zinder captured diverse urban and rural contexts, findings may not be fully generalizable to other regions of Niger characterized by distinct security dynamics or ethnic compositions. Furthermore, reliance on self-reported data introduces the potential for social desirability bias, particularly regarding sensitive topics such as contraceptive use history and hygiene behaviors. Finally, qualitative findings must be interpreted with an awareness of linguistic challenges, as participants explicitly noted difficulties in conceptualizing the term ‘self-care’; consequently, subtle semantic nuances regarding the local distinction between “autonomy” and “soliciting help” may have been attenuated during the translation of interviews from local languages (Haoussa, Zerma) into French.

## Conclusions

The implementation of family planning self-care in Niger requires a departure from a “product-first” approach toward a “values-first” approach. By aligning self-care interventions with local definitions of hygiene, economic survival, and clinical safety, stakeholders can bypass resistance. The path forward lies in a gender-stratified approach: providing “medicalized validation” for women to ensure safety, while leveraging “community-based efficiency” for men to ensure access. Ultimately, self-care in Niger should be framed not as independent from the health system, but as an empowered partnership with it.

## Data Availability

Data are publicly available in the Harvard Dataverse repository at the following URL: https://doi.org/10.7910/DVN/JT3BPZ [17]

## Acknowledgements

We gratefully acknowledge FHI 360, as the lead of the Research for Scalable Solutions (R4S) consortium, for their overarching leadership and support throughout this study. We also extend our sincere appreciation to the consortium partners, EVIHDAF and the Makerere University School of Public Health (MakSPH), for their essential technical and institutional contributions. Special thanks are due to GRADE Africa, who oversaw data collection efforts in Niger. Additionally, we recognize the dedicated efforts of the data management teams at FHI 360, EVIHDAF, and MakSPH for their support in ensuring the integrity and quality of the research findings. Finally, we thank Professor John Cleland, Emeritus Professor at the London School of Hygiene and Tropical Medicine (LSHTM), for his invaluable guidance on the quantitative analyses.

## References

1 World Health Organization. WHO guideline on self-care interventions for health and well-being, 2022 revision. Washington, DC: World Health Organization 2022.

2 Narasimhan M, Logie CH, Gauntley A, et al. Self-care interventions for sexual and reproductive health and rights for advancing universal health coverage. Sexual and Reproductive Health Matters. 2020;28:1778610. doi: 10.1080/26410397.2020.1778610

3 Sundewall J, Williams A, Strauss M, et al. Self-care interventions for sexual and reproductive health: a strategic health systems investment. BMJ Glob Health. 2025;10. doi: 10.1136/bmjgh-2025-019030

4 Borg SA, Narasimhan M, Wuliji T. Mobile learning to support health workers in pharmacies in expanding access to over-the-counter contraceptives: The World Health Organization Academy Learning Programme. Sexual and Reproductive Health Matters. 2022;30:2089084. doi: 10.1080/26410397.2022.2089084

5 Narasimhan M, Aujla M, Lerberghe WV. Self-care interventions and practices as essential approaches to strengthening health-care delivery. The Lancet Global Health. 2023;11:e21–2. doi: 10.1016/S2214-109X(22)00451-X

6 Burke HM, Ridgeway K, Murray K, et al. Reproductive empowerment and contraceptive self-care: a systematic review. Sexual and Reproductive Health Matters. 2022;29:2090057. doi: 10.1080/26410397.2022.2090057

7 Ferguson L, Fried S, Matsaseng T, et al. Human rights and legal dimensions of self care interventions for sexual and reproductive health. BMJ. 2019;365. doi: 10.1136/bmj.l1941

8 High Impact Practices in Family Planning (HIP). Contraceptive Self-Care. Washington, DC: HIP Partnership 2025.

9 Campbell M, Sahin-Hodoglugil NN, Potts M. Barriers to Fertility Regulation: A Review of the Literature. Studies in Family Planning. 2006;37:87–98. doi: 10.1111/j.1728-4465.2006.00088.x

10 Timilsina A, Bhandari B, Johns A, et al. Barriers and facilitators to self-care practices for sexual and reproductive health among women of reproductive age. PLOS ONE. 2024;19:e0303958. doi: 10.1371/journal.pone.0303958

11 Rothschild CW, Sedgh Gi, Brady M, et al. Priority indicators for sexual and reproductive health self-care: recommendations from an expert working group. BMJ Sex Reprod Health. 2023;49:315–6. doi: 10.1136/bmjsrh-2023-201909

12 Logie CH, Berry I, Ferguson L, et al. Uptake and provision of self-care interventions for sexual and reproductive health: findings from a global values and preferences survey. Sexual and Reproductive Health Matters. 2022;29:2009104. doi: 10.1080/26410397.2021.2009104

13 Narasimhan M, Logie CH, Hargreaves J, et al. Self-care interventions for advancing sexual and reproductive health and rights – implementation considerations. Journal of Global Health Reports. 2023;7. doi: 10.29392/001c.84086

14 Logie CH, Khoshnood K, Okumu M, et al. Self care interventions could advance sexual and reproductive health in humanitarian settings. BMJ. 2019;365:l1083. doi: 10.1136/bmj.l1083

15 Jackson A, Angel A, Bagourmé A-RM, et al. A New Contraceptive Diaphragm in Niamey, Niger: A Mixed Methods Study on Acceptability, Use, and Programmatic Considerations. Global Health: Science and Practice. 2022;10. doi: 10.9745/GHSP-D-21-00532

16 Sedgh G, Sorhaindo A. Identifying and prioritizing evidence needs in self-care interventions for sexual and reproductive health. Front Glob Women’s Health. 2023;4. doi: 10.3389/fgwh.2023.1148244

17 Bidashimwa D, Tolley EE, Kedebe K, et al. R4S 1.2. An exploratory, mixed methods study of family planning self-care in Nepal, Niger, and Uganda. 2024. 10.7910/DVN/JT3BPZ

18 Hardon A, Pell C, Taqueban E, et al. Sexual and reproductive self care among women and girls: insights from ethnographic studies. BMJ. 2019;365. doi: 10.1136/bmj.l1333

19 Christofield M, Moon P, Allotey P. Navigating paradox in self-care. BMJ Glob Health. 2021;6. doi: 10.1136/bmjgh-2021-005994

20 Diamond-Smith N, Campbell M, Madan S. Misinformation and fear of side-effects of family planning. *Culture*, Health & Sexuality. 2012;14:421–33.

21 Havaei M, Saeieh SE, Salehi L. Sexual and reproductive health self-care: a theory-based intervention. Health Education. 2021;121:111–24. doi: 10.1108/HE-04-2020-0024

22 Narasimhan M, Logie CH, Moody K, et al. The role of self-care interventions on men’s health-seeking behaviours to advance their sexual and reproductive health and rights. Health Res Policy Sys. 2021;19:23. doi: 10.1186/s12961-020-00655-0

23 Harichund C, Moshabela M. Acceptability of HIV Self-Testing in Sub-Saharan Africa: Scoping Study. AIDS Behav. 2018;22:560–8. doi: 10.1007/s10461-017-1848-9

24 Remme M, Narasimhan M, Wilson D, et al. Self care interventions for sexual and reproductive health and rights: costs, benefits, and financing. BMJ. 2019;365. doi: 10.1136/bmj.l1228

25 Dougherty L, Bellows N, Dadi C. Creating Reproductive Health Behavioral Profiles for Women of Reproductive Age in Niger Using Cross-Sectional Survey Data: A Latent Class Analysis. Int J Public Health. 2023;68:1605247. doi: 10.3389/ijph.2023.1605247

26 Speizer IS, Amani H, Winston J, et al. Assessment of segmentation and targeted counseling on family planning quality of care and client satisfaction: a facility-based survey of clients in Niger. BMC Health Serv Res. 2021;21:1075. doi: 10.1186/s12913-021-07066-z

27 Brady M, Drake JK, Namagembe A, et al. Self-care provision of contraception: Evidence and insights from contraceptive injectable self-administration. Best Practice & Research Clinical Obstetrics & Gynaecology. 2020;66:95–106. doi: 10.1016/j.bpobgyn.2020.01.003

28 Makusha T, Knight L, Taegtmeyer M, et al. HIV Self-Testing Could “Revolutionize Testing in South Africa, but It Has Got to Be Done Properly”: Perceptions of Key Stakeholders. PLOS ONE. 2015;10:e0122783. doi: 10.1371/journal.pone.0122783

29 Nouhou A-M, Oumara MSH, Assoumane AI, et al. Transforming and integrating gender norms and social practices to promote maternal health through male engagement. Afr J Reprod Health. 2025;29:76–88. doi: 10.29063/ajrh2025/v29i6s.6

30 Di Giorgio L, Mvundura M, Tumusiime J, et al. Costs of administering injectable contraceptives through health workers and self-injection: evidence from Burkina Faso, Uganda, and Senegal. CONTRACEPTION. 2018;98:389–95. doi: 10.1016/j.contraception.2018.05.018

31 Burke H, Mueller M, Packer C, et al. Provider acceptability of Sayana® Press: results from communityA health workers and clinic-based providers in Uganda and Senegal. CONTRACEPTION. 2014;89:368–73. doi: 10.1016/j.contraception.2014.01.009

32 Cover J, Ba M, Lim J, et al. Evaluating the feasibility and acceptability of self-injection of subcutaneous depot medroxyprogesterone acetate (DMPA) in Senegal: a prospective cohort study. CONTRACEPTION. 2017;96:203–10. doi: 10.1016/j.contraception.2017.06.010

33 Kennedy CE, Yeh PT, Gonsalves L, et al. Should oral contraceptive pills be available without a prescription? A systematic review of over-the-counter and pharmacy access availability. BMJ Glob Health. 2019;4. doi: 10.1136/bmjgh-2019-001402

34 Stout A, Wood S, Barigye G, et al. Expanding Access to Injectable Contraception: Results From Pilot Introduction of Subcutaneous Depot Medroxyprogesterone Acetate (DMPA-SC) in 4 African Countries. Global Health: Science and Practice. 2018;6:55–72. doi: 10.9745/GHSP-D-17-00250

35 Institut National de la Statistique (INS) et Utica International. Enquête Nationale sur la Fécondité et la Mortalité des Enfants de Moins de Cinq Ans au Niger 2021. Niamey, Niger et Columbia, Maryland, USA: INS et Utica International 2022.

36 Brittingham S, Mitchell L, Zan T. What’s inside matters: an assessment of the family planning content of digital self-care platforms. Reprod Health. 2024;21:112. doi: 10.1186/s12978-024-01848-4

